# One-year efficacy and tolerability of 0.05% atropine for myopia control in Estonia: a prospective cohort study

**DOI:** 10.64898/2026.04.02.26348423

**Authors:** Delis Linntam, Kadi Palumaa, Teele Palumaa

**Author notes:** **Corresponding author:** Teele Palumaa, MD, PhD.

## Abstract

**Background:** Despite strong evidence from controlled trials, uncertainty remains about the real-world use of 0.05% atropine in patients with lighter irises due to tolerability concerns, and predictors of treatment response are poorly understood. Here, we evaluated the effectiveness, tolerability, and early biometric response to 0.05% atropine in clinical practice among patients with predominantly light irises.

**Methods:** This prospective cohort study included 33 patients treated with 0.05% atropine (82% with light irises). Cycloplegic spherical equivalent refraction (SER) was measured at baseline and 3-month intervals. Axial length (AL), photopic pupil diameter, accommodation amplitude, and subjective side effects were monitored more frequently initially.

**Results:** Median age at treatment initiation was 11.97 years, SER −5.38 D, and AL 25.42 mm. Over 12 months, SER changed by –0.078 ± 0.349 D (mean ± SD), and AL increased by 0.052 ± 0.115 mm. Eighty-eight percent of participants had a SER change of <0.5 D, and 91% had axial elongation of <0.2 mm, indicating clinically limited myopia progression. Photopic pupil diameter was larger, and accommodation amplitude was reduced throughout follow-up. Early in treatment, side effects, including photophobia and near-work difficulties, were common but minimally disruptive. Their incidence decreased rapidly and rarely required treatment modification. In exploratory analyses, early AL changes predicted 12-month AL outcomes, with associations detectable as early as 1 week and strengthening over time.

**Conclusions:** 0.05% atropine was well tolerated and effective in this population with light irises. Early AL changes may predict 12-month treatment response. These findings support the implementation of 0.05% atropine in routine clinical practice in populations with light irises and highlight the potential for early AL monitoring to guide timely treatment adjustments.

## Background

The prevalence of myopia has increased rapidly over the last decades, and it is estimated that by the year 2050, approximately 40-50% of the global population will be affected [1, 2]. Although refractive correction with spectacles or contact lenses is effective, myopia is a major risk factor for several potentially sight-threatening conditions later in life, including myopic macular degeneration, retinal detachment, glaucoma, and cataract [3–5]. The risk increases with myopia severity and age, and is associated with a higher likelihood of irreversible visual impairment [6]. Slowing the progression of myopia in children and adolescents is therefore a high clinical priority.

Topical atropine is one of the most effective interventions for myopia control [7–9]. Clinical studies have shown that low-concentration atropine slows myopia progression in a dose-dependent manner, with higher concentrations providing greater efficacy [10–12], but also a higher incidence of side effects [10–14]. Among low-concentration atropine, 0.05% offers the most favourable efficacy, and has therefore been increasingly adopted in clinical practice as a first-line pharmacological therapy for progressive myopia [11, 12].

However, side effects remain a concern: separate clinical trials report that almost a third of patients require photochromic glasses [15], or 15% experience photophobia [16]. Tolerability profile has received particular attention in populations with light irises. Iris pigmentation affects the susceptibility of the pupil to atropine-induced dilation [17], which could underlie the higher incidence of atropine-related side effects in those with lighter irises. Indeed, individuals with light irises experience greater pupil dilation when treated with 0.01% atropine.[18] In real-world settings with mixed iris pigmentation, up to 63% of patients reported near-work difficulties shortly after treatment initiation [14], raising concerns about the feasibility of using 0.05% atropine in routine clinical practice [19].

Beyond tolerability, another key challenge in atropine therapy is identifying early indicators of treatment response. From a clinical perspective, tools to predict long-term treatment response early in therapy would be valuable for tailoring management strategies; however, no established methods currently exist to guide timely decision-making.

In this study, we report the 12-month treatment effect and tolerability profile of 0.05% atropine in 33 patients with predominantly light irises treated in routine clinical practice. In addition, we explore whether early biometric changes could predict long-term treatment response.

## Methods

### Ethics

The study was approved by the Tallinn Medical Research Ethics Committee and adhered to the tenets of the Declaration of Helsinki. Written informed consent was obtained from a parent or guardian of participants younger than 18 years and from participants older than 12 years; oral assent was obtained from participants younger than 12 years.

### Study population

Eligible participants met the East Tallinn Central Hospital criteria for atropine treatment, defined as baseline axial length (AL) ≥75th percentile on the Dutch population growth chart [20] and/or myopia progression ≥0.50 dioptres (D) per year. In participants with unilateral myopia requiring treatment (n = 2), only the treated eye was analysed. In one participant with a history of congenital cataract surgery and pseudophakia in one eye, analyses were restricted to the fellow phakic eye. The final study sample comprised 33 participants.

### Intervention

0.05% atropine eye drops were compounded as preservative-free, single-dose units at the East Tallinn Central Hospital pharmacy. In accordance with Estonian regulations, five units were dispensed weekly. Patients and their parents/guardians were instructed to instil one drop in each eye every evening, Monday to Friday. Two participants did not initially tolerate 0.05% atropine and were temporarily switched to a lower concentration (0.025% or 0.01%); both resumed 0.05% atropine by 6 and 9 months, respectively, and were retained in the primary analysis.

### Outcome measures

Participants underwent a baseline visit and follow-up assessments at 1 week, 1 month, and 3, 6, 9, and 12 months. AL, photopic pupil diameter, and accommodation amplitude were measured at each visit. AL and pupil diameter were measured using the Topcon MYAH, which has demonstrated good repeatability, reproducibility and accuracy comparable to that of stand-alone biometers [21]. Accommodation amplitude was measured monocularly using a RAF rule. Anterior segment examination was performed at each visit using slit-lamp biomicroscopy to assess any pathologies. Eye colour was graded at baseline using the Seddon scale, which rates pigmentation on a four-point scale from light to dark [22].

Cycloplegic spherical equivalent refraction (SER) was assessed at baseline and at 3-month intervals thereafter. Cycloplegia was induced using two drops of 1% cyclopentolate administered 5 minutes apart. Autorefraction was performed with the Nidek HandyRef-K 40 minutes after the first instillation. Best-corrected visual acuity (BCVA) was measured after cycloplegia using Snellen charts, with pinhole testing if acuity was <0.9 decimal. Dilated fundus examination was conducted to identify ocular findings associated with myopia or other pathology. No new abnormalities were detected on anterior segment or fundus biomicroscopy during follow-up.

At each follow-up visit, subjective side effects were systematically recorded using a structured checklist. Participants were asked whether they experienced enlarged pupils, light sensitivity, difficulty with near work, eye redness, irritation, headaches, or any other symptoms. For each reported side effect, participants were asked whether the symptom caused any disturbance in daily activities and to rate the severity of the symptom on a 0–10 scale (0 = no disturbance, 10 = greatest disturbance imaginable).

### Statistical analyses

Data were collected in REDCap and analysed in de-identified form. Statistical analyses and visualisation were performed using R (version 4.3.3; R Foundation for Statistical Computing, Vienna, Austria). Continuous normally distributed variables were summarised as mean and standard deviation (SD), and non-normally distributed variables as median and interquartile range [IQR] or first and third quartiles [Q1, Q3].

For analyses involving ocular measurements, values were averaged across both eyes unless only one eye was available (see the Study population section). Changes in ocular parameters were calculated as the difference between baseline and follow-up visits. Longitudinal changes in cycloplegic SER, AL, pupil size, and accommodation amplitude were analysed using linear mixed-effects models with fixed effects for time point, age group (<12 years vs ≥12 years), and their interaction, and a random intercept for participant; baseline served as the reference time point.

To assess whether early biometric changes predicted treatment response at 12 months, linear regression analyses were performed with AL change from baseline to 12 months as the outcome. Separate models were fitted using AL change at 1 week, 1 month, 3 months, and 6 months as predictors. All models were adjusted for baseline AL, age, and sex. Model performance was summarised using adjusted R^2^. A *P* value <0.05 was considered statistically significant.

## Results

The study included 33 participants with a median age of 11.97 years (Q1, Q3: 9.81, 12.74, range: 4.65, 18.0) and a balanced sex distribution (48.5% male). At baseline, participants had moderate to high myopia, with a median SER of −5.38 D (Q1, Q3: −7.50, −2.81, range: −11.5 to −0.625) and a median axial length of 25.42 mm (Q1, Q3: 24.16, 26.05, range: 23.04 to 28.47). Baseline characteristics of the cohort, overall and stratified by age group (<12 years, ≥12 years), are shown in **Table 1**.

**Table 1.**
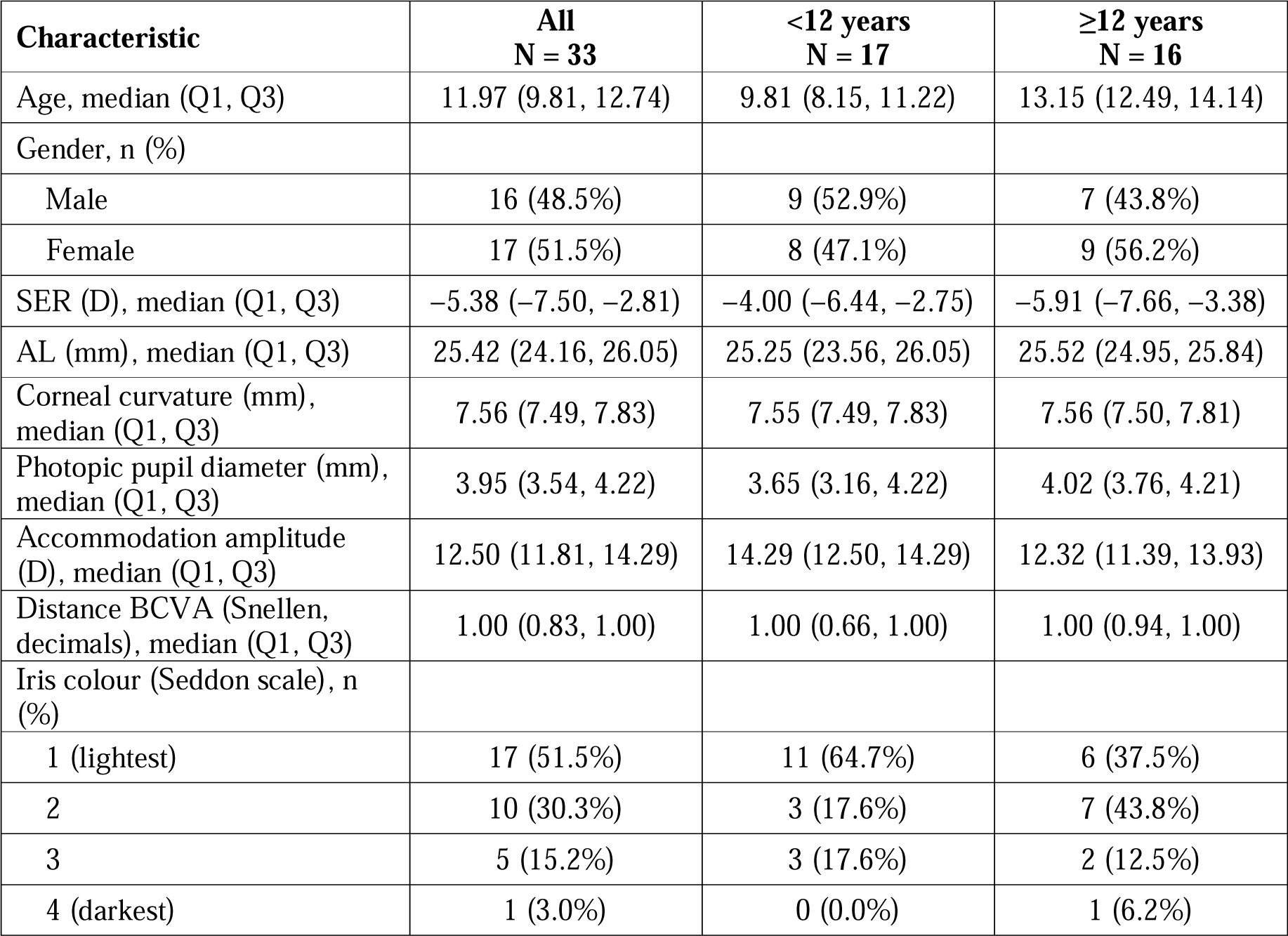
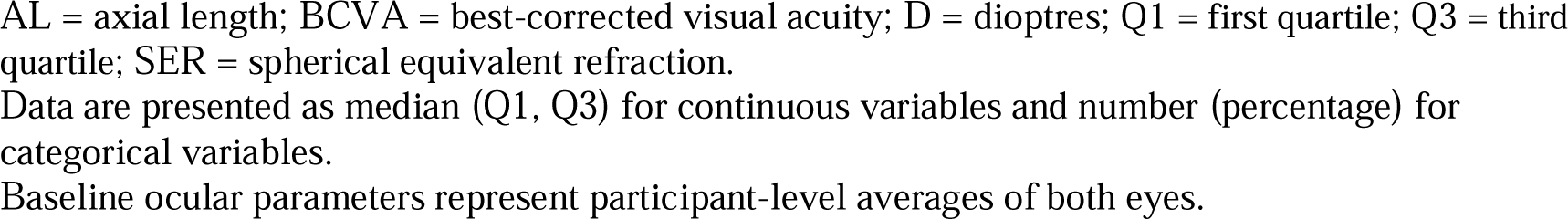
Baseline demographic and ocular characteristics of the study participants.

| Characteristic | All<br>N = 33 | <12 years<br>N = 17 | ≥12 years<br>N = 16 |
| --- | --- | --- | --- |
| Age, median (Q1, Q3) | 11.97 (9.81, 12.74) | 9.81 (8.15, 11.22) | 13.15 (12.49, 14.14) |
| Gender, n (%) |  |  |  |
| Male | 16 (48.5%) | 9 (52.9%) | 7 (43.8%) |
| Female | 17 (51.5%) | 8 (47.1%) | 9 (56.2%) |
| SER (D), median (Q1, Q3) | -5.38 (-7.50, -2.81) | -4.00 (-6.44, -2.75) | -5.91 (-7.66, -3.38) |
| AL (mm), median (Q1, Q3) | 25.42 (24.16, 26.05) | 25.25 (23.56, 26.05) | 25.52 (24.95, 25.84) |
| Corneal curvature (mm), median (Q1, Q3) | 7.56 (7.49, 7.83) | 7.55 (7.49, 7.83) | 7.56 (7.50, 7.81) |
| Photopic pupil diameter (mm), median (Q1, Q3) | 3.95 (3.54, 4.22) | 3.65 (3.16, 4.22) | 4.02 (3.76, 4.21) |
| Accommodation amplitude (D), median (Q1, Q3) | 12.50 (11.81, 14.29) | 14.29 (12.50, 14.29) | 12.32 (11.39, 13.93) |
| Distance BCVA (Snellen, decimals), median (Q1, Q3) | 1.00 (0.83, 1.00) | 1.00 (0.66, 1.00) | 1.00 (0.94, 1.00) |
| Iris colour (Seddon scale), n (%) |  |  |  |
| 1 (lightest) | 17 (51.5%) | 11 (64.7%) | 6 (37.5%) |
| 2 | 10 (30.3%) | 3 (17.6%) | 7 (43.8%) |
| 3 | 5 (15.2%) | 3 (17.6%) | 2 (12.5%) |
| 4 (darkest) | 1 (3.0%) | 0 (0.0%) | 1 (6.2%) |

AL = axial length; BCVA = best-corrected visual acuity; D = dioptres; Q1 = first quartile; Q3 = third quartile; SER = spherical equivalent refraction.
Data are presented as median (Q1, Q3) for continuous variables and number (percentage) for categorical variables.
Baseline ocular parameters represent participant-level averages of both eyes.

After 12 months of treatment, the mean change in cycloplegic SER was −0.078 ± 0.349 D (mean ± SD), and 87.9% of participants exhibited a SER change of <0.5 D (**Table 2**, **Figure 1a-b**). Axial length increased on average by 0.052 ± 0.115 mm, and 90.9% had an AL change of <0.2 mm/year (**Figure 1c-d**). Accommodation amplitude decreased early during treatment and remained reduced throughout follow-up, while photopic pupil diameter increased and remained enlarged over time. No significant differences in longitudinal outcomes were observed between age groups (**Table 2**).

**Figure 1.**
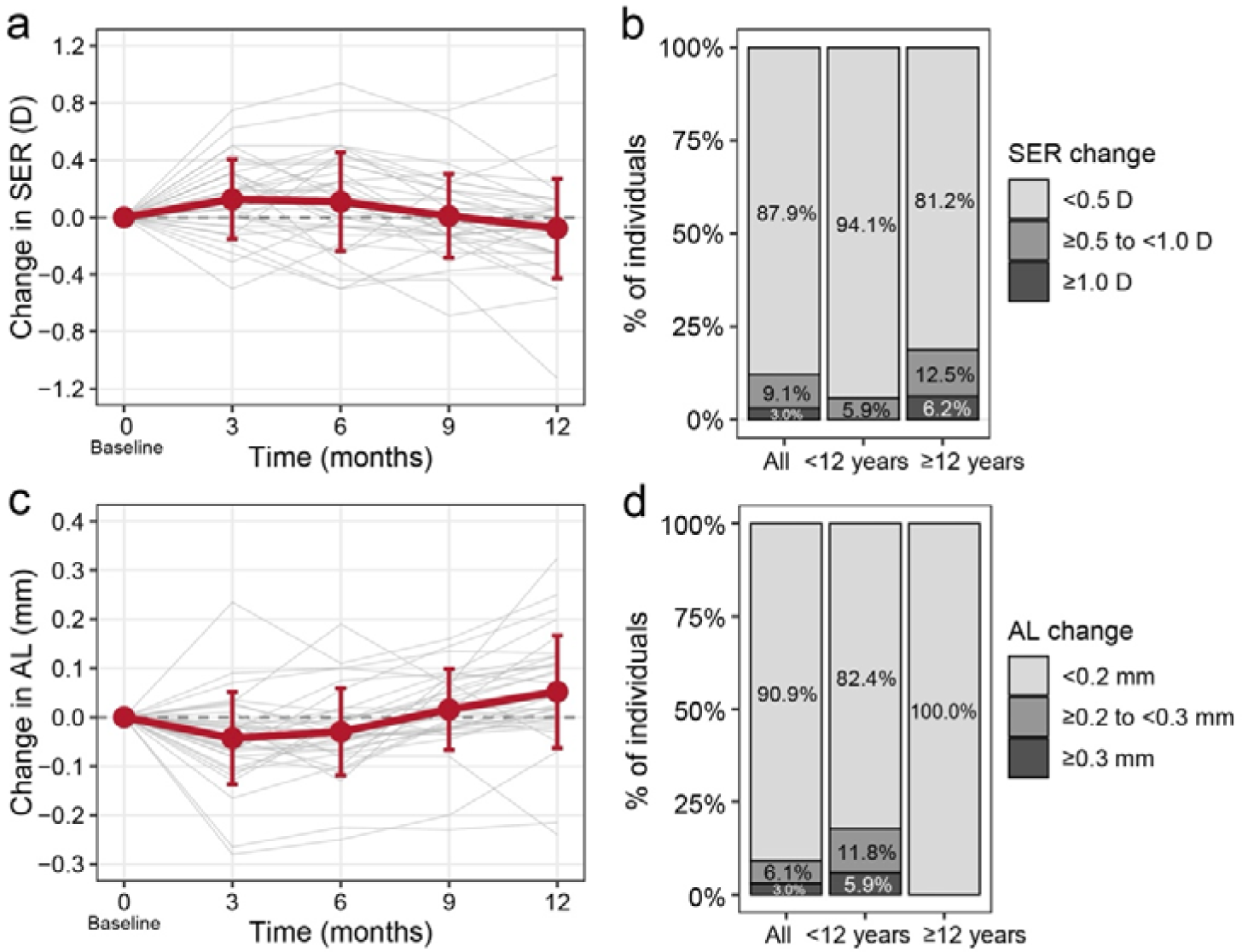
Longitudinal changes in spherical equivalent and axial length, and distribution of treatment response. (a) Change in cycloplegic spherical equivalent refraction (SER) from baseline across follow-up visits. (b) Distribution of treatment response at 12 months based on SER change from baseline. (c) Change in AL from baseline across follow-up visits. (d) Distribution of treatment response at 12 months based on AL elongation from baseline. Panels (a) and (c) depict mean ± SD; individual participants are shown in grey. AL = axial length; D = dioptres; SD = standard deviation; SER = spherical equivalent refraction.

**Table 2.** Longitudinal changes in ocular parameters over 12 months.

|  | All |  | <12 years |  | ≥12 years |  | P value<br>(12 m vs<br>baseline) | P value<br>(age group<br>× time) |
| --- | --- | --- | --- | --- | --- | --- | --- | --- |
| Change | Mean | SD | Mean | SD | Mean | SD |  |  |
| Spherical equivalent refraction (D) |  |  |  |  |  |  |  |  |
| Baseline to 3 months | 0.127 | 0.278 | 0.160 | 0.285 | 0.094 | 0.276 | 0.35 | 0.29 |
| Baseline to 6 months | 0.110 | 0.345 | 0.096 | 0.329 | 0.125 | 0.372 |  |  |
| Baseline to 9 months | 0.009 | 0.294 | -0.026 | 0.316 | 0.047 | 0.272 |  |  |
| Baseline to 12 months | -0.078 | 0.349 | -0.015 | 0.346 | -0.145 | 0.350 |  |  |
| Axial length (mm) |  |  |  |  |  |  |  |  |
| Baseline to 1 week | -0.026 | 0.069 | -0.028 | 0.074 | -0.024 | 0.065 | 0.004 | 0.12 |
| Baseline to 1 month | -0.018 | 0.064 | 0.000 | 0.048 | -0.038 | 0.074 |  |  |
| Baseline to 3 months | -0.043 | 0.094 | -0.029 | 0.090 | -0.057 | 0.100 |  |  |
| Baseline to 6 months | -0.030 | 0.089 | -0.006 | 0.086 | -0.055 | 0.087 |  |  |
| Baseline to 9 months | 0.016 | 0.083 | 0.036 | 0.072 | -0.006 | 0.090 |  |  |
| Baseline to 12 months | 0.052 | 0.115 | 0.088 | 0.128 | 0.013 | 0.087 |  |  |
| Accommodation amplitude (D) |  |  |  |  |  |  |  |  |
| Baseline to 1 week | -2.87 | 3.33 | -3.09 | 2.84 | -2.65 | 3.84 | 0.011 | 0.57 |
| Baseline to 1 month | -2.26 | 3.21 | -1.67 | 3.62 | -2.93 | 2.62 |  |  |
| Baseline to 3 months | -1.74 | 2.75 | -1.58 | 3.53 | -1.89 | 1.76 |  |  |
| Baseline to 6 months | -1.56 | 3.29 | -1.27 | 4.39 | -1.88 | 1.53 |  |  |
| Baseline to 9 months | -0.95 | 2.63 | -0.92 | 3.49 | -0.98 | 1.33 |  |  |
| Baseline to 12 months | -1.97 | 2.61 | -2.56 | 2.94 | -1.37 | 2.17 |  |  |
| Photopic pupil diameter (mm) |  |  |  |  |  |  |  |  |
| Baseline to 1 week | 1.16 | 0.89 | 0.98 | 0.90 | 1.34 | 0.88 | <0.001 | 0.62 |
| Baseline to 1 month | 0.85 | 0.61 | 0.75 | 0.65 | 0.93 | 0.59 |  |  |
| Baseline to 3 months | 0.78 | 0.69 | 0.70 | 0.88 | 0.87 | 0.40 |  |  |
| Baseline to 6 months | 0.99 | 0.87 | 0.96 | 0.81 | 1.02 | 0.95 |  |  |
| Baseline to 9 months | 0.80 | 0.84 | 0.79 | 0.78 | 0.82 | 0.93 |  |  |
| Baseline to 12 months | 0.97 | 0.94 | 1.08 | 0.86 | 0.86 | 1.02 |  |  |
| Distance BCVA (Snellen, decimals) |  |  |  |  |  |  |  |  |
| Baseline to 3 months | -0.02 | 0.23 | 0.02 | 0.16 | -0.06 | 0.28 | 0.95 | 0.45 |
| Baseline to 6 months | -0.01 | 0.21 | 0.02 | 0.10 | -0.04 | 0.27 |  |  |
| Baseline to 9 months | -0.03 | 0.20 | 0.01 | 0.12 | -0.07 | 0.25 |  |  |
| Baseline to 12 months | 0.01 | 0.22 | 0.05 | 0.13 | -0.03 | 0.27 |  |  |

BCVA = best-corrected visual acuity; D = dioptres; SD = standard deviation.
Values represent change from baseline using participant-level averages of both eyes.
*P* values for baseline to 12-month comparisons were derived from linear mixed-effects models including timepoint, age group, and their interaction, with participant as a random effect.
*P* values for the age group × time interaction represent a global test of whether longitudinal trajectories differed between age groups across follow-up.

Ocular side effects were common shortly after treatment initiation and decreased rapidly over time. Enlarged pupils were reported by 69.7% of participants at 1 week, declining to 24.2% at 3 months and 12.1% at 6 months (**Table 3**). Photophobia and difficulty with near work affected 48.5% and 42.4% of participants at 1 week, respectively, and 12.1% at 3 months. Median severity scores for these symptoms decreased from 4–5 on a 10-point scale at 1 week to 2 at 3 months, indicating limited interference with daily activities. Eye redness and irritation affected fewer than 10% of participants at any time point, and no systemic side effects were reported. Two participants experienced severe near-work difficulties that interfered with schoolwork early in treatment and were switched to lower concentrations (0.025% or 0.01%). Both subsequently resumed 0.05% atropine during follow-up at 6 and 9 months, respectively. No participants required photochromic or progressive glasses, and none discontinued treatment.

**Table 3.** Frequency and severity of side effects over 12 months.

| Side effect | Measure | 1 week | 1 month | 3 months | 6 months | 9 months | 12 months |
| --- | --- | --- | --- | --- | --- | --- | --- |
| Enlarged pupils | n (%) | 23 (69.7%) | 16 (48.5%) | 8 (24.2%) | 4 (12.1%) | 3 (9.1%) | 3 (9.1%) |
|  | Severity score, median (Q1, Q3) | 4 (2.2, 5.0) | 3 (2.0, 3.5) | 2 (0.0, 4.0) | 0 (0.0, 2.5) | 2 (1.0, 2.5) | 0 (0.0, 0.0) |
| Photophobia | n (%) | 16 (48.5%) | 11 (33.3%) | 4 (12.1%) | 0 (0.0%) | 0 (0.0%) | 3 (9.1%) |
|  | Severity score, median (Q1, Q3) | 5 (2.0, 5.0) | 2 (2.0, 3.0) | 2 (2.0, 2.0) | — | — | 3 (3.0, 3.5) |
| Near-work difficulties | n (%) | 14 (42.4%) | 6 (18.2%) | 4 (12.1%) | 0 (0.0%) | 3 (9.1%) | 0 (0.0%) |
|  | Severity score, median (Q1, Q3) | 4 (3.0, 5.0) | 3 (0.8, 3.8) | 2 (1.0, 2.0) | — | 2 (1.0, 2.5) | — |
| Eye redness | n (%) | 2 (6.1%) | 1 (3.0%) | 2 (6.1%) | 1 (3.0%) | 1 (3.0%) | 0 (0.0%) |
|  | Severity score, median (Q1, Q3) | 1 (1.0, 1.0) | NA | 5 | 5 | 3 | — |
| Eye irritation | n (%) | 1 (3.0%) | 0 (0.0%) | 1 (3.0%) | 1 (3.0%) | 0 (0.0%) | 0 (0.0%) |
|  | Severity score, median (Q1, Q3) | NA | — | 5 | 5 | — | — |
| Headache | n (%) | 0 (0.0%) | 0 (0%) | 0 (0%) | 0 (0%) | 0 (0%) | 0 (0%) |
|  | Severity score, median (Q1, Q3) | — | — | — | — | — | — |

Q1 = first quartile; Q3 = third quartile.
Data are presented as number (percentage) of participants reporting each side effect at each visit.
All 33 participants were systematically assessed for the listed side effects at each follow-up visit.
Severity scores are summarised as median (Q1, Q3) when applicable.
NA indicates that the adverse event was reported but the severity grade was not recorded; — indicates that the adverse event did not occur.

Interestingly, we observed a transient increase in SER and a decrease in AL during the first 6 months of follow-up (**Table 2**, **Figure 1a, c**), which prompted us to explore whether early AL changes were associated with 12-month AL outcomes. In exploratory regression analyses adjusted for age, sex, and baseline AL, greater early AL elongation (at 1 week, 1 month, 3 months, and 6 months) was consistently associated with greater AL elongation at 12 months (*P* < 0.05 for all; **Table 4**, **Figure 2**). The model’s explanatory power was smallest at 1 week and increased at later time points, with a strong association at 3 months (adjusted R^2^ = 0.40; *P* = 0.0025; **Figure 2a**) and the strongest at 6 months (adjusted R^2^ = 0.66; *P* < 0.001; **Figure 2b**). At 6 months, the association approached a 1:1 relationship (β = 0.95), suggesting that early AL change closely tracks long-term outcome.

**Figure 2.**
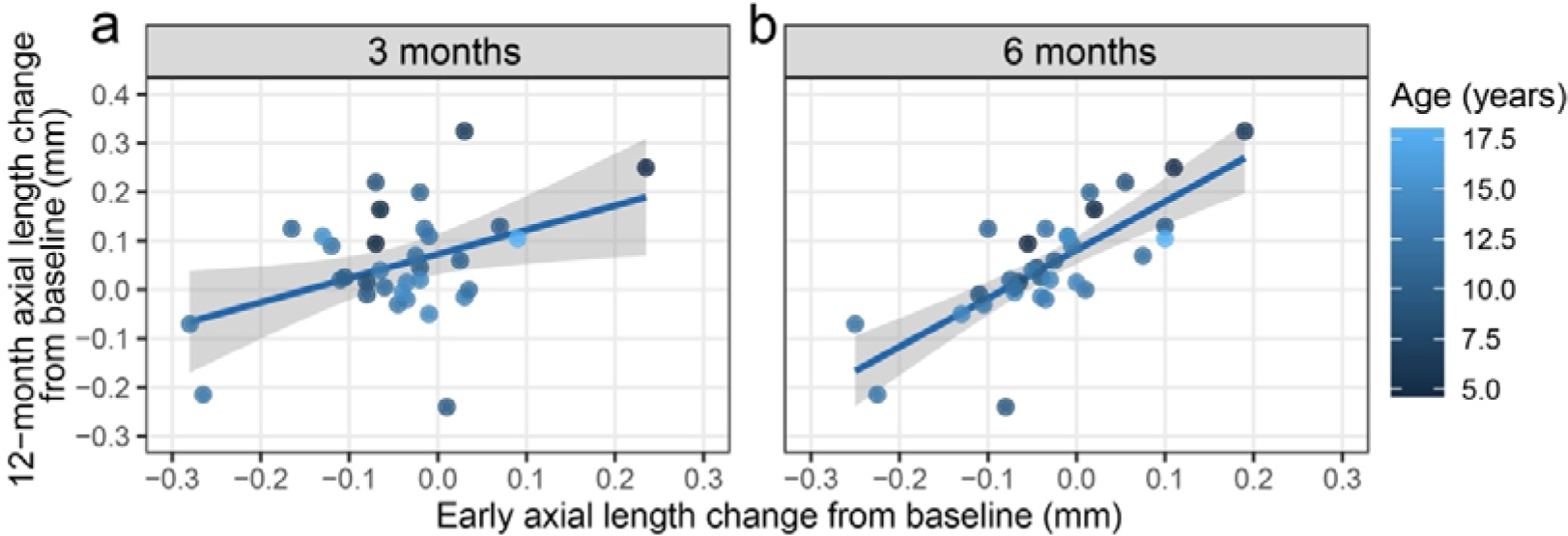
Relationship between early axial length change and 12-month outcome. Associations between axial length (AL) change at 3 months (a) and 6 months (b) and AL change at 12 months. Each point represents one participant; solid lines indicate linear regression with 95% confidence intervals. Colour indicates age at treatment initiation.

**Table 4.** Association between early axial length change and 12-month axial length outcome.

| Predictor | $\beta$ (95% CI) | *P* value | Adjusted $R^2$ |
| --- | --- | --- | --- |
| ΔAL at 1 week | 0.55 (0.001–1.108) | 0.0496 | 0.27 |
| ΔAL at 1 month | 0.87 (0.263–1.472) | 0.0065 | 0.36 |
| ΔAL at 3 months | 0.66 (0.251–1.060) | 0.0025 | 0.40 |
| ΔAL at 6 months | 0.95 (0.651–1.254) | <0.001 | 0.66 |

AL = axial length; CI = confidence interval.
$\beta$ coefficients and 95% CIs were derived from linear regression models adjusted for age, sex, and baseline axial length.
$\Delta$ AL represents change in axial length relative to baseline at the specified time point.
Adjusted $R^2$ indicates the proportion of variance in 12-month axial length change explained by the full model.

## Discussion

In this prospective cohort study, 0.05% atropine treatment was associated with minimal SER progression (–0.078 ± 0.349 D) and a very modest axial elongation (0.052 ± 0.115 mm) over one year. Although ocular side effects were frequently reported shortly after treatment initiation, their rapid attenuation over time indicates good overall tolerability. Taken together, these observations support the clinical feasibility of 0.05% atropine for myopia control in routine practice among patients with light irises.

The magnitude of treatment effect observed in this cohort was broadly comparable to that reported in other studies using 0.05% atropine. The MOSAIC2 study, conducted in Ireland in a slightly older cohort (mean age 13.7 years), reported changes of –0.11 D in SER and 0.09 mm in AL over one year [12]. In the LAMP study, which included a younger population (mean age 8.5 years), the one-year change in SER was –0.27 D and AL 0.20 mm [15]. Age is a well-established modifier of treatment response, with young children typically demonstrating faster myopia progression and smaller treatment response to atropine [23]. Iris pigmentation may also influence both the efficacy and tolerability of atropine treatment. For example, in the MOSAIC study, 0.01% atropine demonstrated a treatment effect in those with blue irises, but not in those with green or brown irises [16]. The excellent treatment response in this cohort may partly reflect the participants’ relatively older age and light irises. Slower progression rates observed in real-world settings compared with clinical trials may also have contributed [24].

The side-effect profile in this cohort provides reassurance about tolerability in routine clinical practice for patients with light irises, who have been reported in some studies to experience more side effects [18]. Similar to studies assessing short-term effects of 0.05% atropine [14, 25], we observed a high incidence of ocular side effects early in treatment, most commonly enlarged pupils and photophobia. Importantly, the incidence declined rapidly over time. Furthermore, although early side effects were frequent, their severity was generally mild. Regarding side effects later in treatment, these were reported in a small fraction of patients. Only 9% reported photophobia or near-work difficulties beyond 6 months. These are comparable to the MOSAIC2 study, in which blurred near vision and photophobia were reported by 15% and 8% of participants, respectively [12]. In the LAMP study, photophobia was more common and 31% of patients needed photochromic glasses [15]. Taken together, these findings suggest that while side effects with 0.05% atropine are common early in treatment, they are typically transient and rarely require adjustment in a real-world clinical setting.

In our exploratory analysis, early changes in axial length were associated with axial elongation at 12 months, with associations observed as early as 1 week after treatment initiation and strengthening at later time points (**Table 4**). As expected, the adjusted regression model incorporating axial length change at 6 months explained the largest proportion of variance in the 12-month outcome; however, the presence of detectable associations at 1 week, 1 month, and 3 months is noteworthy. A similar observation has been reported for repeated low-level red-light therapy, in which the 3-month change in AL was a good predictor of treatment response at 12 and 24 months [26]. In a study on orthokeratology, axial elongation over the first treatment year was associated with treatment effect at six years [27]. Although our findings are limited by sample size and should be interpreted as hypothesis-generating, they suggest that early biometric response to atropine may provide clinically useful information. If confirmed in larger cohorts, these early indicators could help identify children with suboptimal treatment response and support timely adjustments, such as dose escalation or combination therapy, rather than waiting for continued progression to become apparent.

This study has several limitations. First, the sample size was modest, limiting statistical power for subgroup analyses. Second, the absence of an untreated control group in this real-world study makes it difficult to distinguish treatment effects from the normal course of eye growth in this population. Third, follow-up was limited to 12 months, precluding assessment of longer-term efficacy and potential rebound effects. Finally, the exploratory regression models should be interpreted cautiously, as no external validation was performed.

## Conclusions

Our findings suggest that 0.05% atropine is a feasible and well-tolerated option for myopia control in real-world clinical settings, including in populations with predominantly light irises. Larger studies with longer follow-up are warranted to validate these observations, refine strategies for personalised myopia management, and develop tools for early identification of treatment responders.

## Declarations

### Ethics approval and consent to participate

The study was approved by the Tallinn Medical Research Ethics Committee and adhered to the tenets of the Declaration of Helsinki. Written informed consent was obtained from a parent or guardian of participants younger than 18 years and from participants older than 12 years; oral assent was obtained from participants younger than 12 years.

### Consent for publication

Not applicable

### Availability of data and materials

All data generated or analysed during this study are included in this published article.

### Authors’ contributions

DL, KP, and TP conceptualised and designed the study. DL, KP, and TP acquired the data. DL and TP analysed the data. DL and TP wrote the first draft of the manuscript. TP supervised the study. DL, KP, and TP reviewed and approved the final manuscript.

## Data Availability

All data produced in the present work are contained in the manuscript

## Acknowledgements

The authors thank the East Tallinn Central Hospital pharmacy staff, particularly Ülle Meren and Jekaterina Semennikova, for introducing the compounding of atropine eye drops at the hospital. We also thank the staff at the East Tallinn Central Hospital Eye Clinic, especially Dr Katrin Hannus, for organising atropine dispensing logistics, and Terje Peetersoo and Kerttu Michelson for their assistance in dispensing the medication to patients.

## List of abbreviations

AL: axial length
BCVA: best-corrected visual acuity
D: dioptre
IQR: interquartile range
Q1: first quartile
Q3: third quartile
SD: standard deviation
SER: spherical equivalent refraction

## References

1. Holden BA, Fricke TR, Wilson DA, Jong M, Naidoo KS, Sankaridurg P, et al. Global Prevalence of Myopia and High Myopia and Temporal Trends from 2000 through 2050. Ophthalmology. 2016;123:1036–42. 10.1016/j.ophtha.2016.01.006.

2. Liang J, Pu Y, Chen J, Liu M, Ouyang B, Jin Z, et al. Global prevalence, trend and projection of myopia in children and adolescents from 1990 to 2050: a comprehensive systematic review and meta-analysis. Br J Ophthalmol. 2025;109:362–71. 10.1136/bjo-2024-325427.

3. Flitcroft DI. The complex interactions of retinal, optical and environmental factors in myopia aetiology. Prog Retin Eye Res. 2012;31:622–60. 10.1016/j.preteyeres.2012.06.004.

4. Wong Y-L, Sabanayagam C, Ding Y, Wong C-W, Yeo AC-H, Cheung Y-B, et al. Prevalence, Risk Factors, and Impact of Myopic Macular Degeneration on Visual Impairment and Functioning Among Adults in Singapore. Investig Opthalmology Vis Sci. 2018;59:4603. 10.1167/iovs.18-24032.

5. Haarman AEG, Enthoven CA, Tideman JWL, Tedja MS, Verhoeven VJM, Klaver CCW. The Complications of Myopia: A Review and Meta-Analysis. Invest Ophthalmol Vis Sci. 2020;61:49. 10.1167/iovs.61.4.49.

6. Tideman JWL, Snabel MCC, Tedja MS, Van Rijn GA, Wong KT, Kuijpers RWAM, et al. Association of Axial Length With Risk of Uncorrectable Visual Impairment for Europeans With Myopia. JAMA Ophthalmol. 2016;134:1355. 10.1001/jamaophthalmol.2016.4009.

7. Chua W-H, Balakrishnan V, Chan Y-H, Tong L, Ling Y, Quah B-L, et al. Atropine for the Treatment of Childhood Myopia. Ophthalmology. 2006;113:2285–91. 10.1016/j.ophtha.2006.05.062.

8. Wildsoet CF, Chia A, Cho P, Guggenheim JA, Polling JR, Read S, et al. IMI – Interventions for Controlling Myopia Onset and Progression Report. Invest Ophthalmol Vis Sci. 2019;60:M106–31. 10.1167/iovs.18-25958.

9. Lawrenson JG, Huntjens B, Virgili G, Ng S, Dhakal R, Downie LE, et al. Interventions for myopia control in children: a living systematic review and network meta[analysis. Cochrane Database Syst Rev. 2025. 10.1002/14651858.CD014758.pub3.

10. Chia A, Chua W-H, Cheung Y-B, Wong W-L, Lingham A, Fong A, et al. Atropine for the Treatment of Childhood Myopia: Safety and Efficacy of 0.5%, 0.1%, and 0.01% Doses (Atropine for the Treatment of Myopia 2). Ophthalmology. 2012;119:347–54. 10.1016/j.ophtha.2011.07.031.

11. Yam JC, Zhang XJ, Zhang Y, Yip BHK, Tang F, Wong ES, et al. Effect of Low-Concentration Atropine Eyedrops vs Placebo on Myopia Incidence in Children: The LAMP2 Randomized Clinical Trial. JAMA. 2023;329:472. 10.1001/jama.2022.24162.

12. Loughman J, Lingham G, Nkansah EK, Kobia-Acquah E, Flitcroft DI. Efficacy and Safety of Different Atropine Regimens for the Treatment of Myopia in Children: Three-Year Results of the MOSAIC Randomized Clinical Trial. JAMA Ophthalmol. 2025;143:134–44. 10.1001/jamaophthalmol.2024.5703.

13. Gong Q, Janowski M, Luo M, Wei H, Chen B, Yang G, et al. Efficacy and Adverse Effects of Atropine in Childhood Myopia: A Meta-analysis. JAMA Ophthalmol. 2017;135:624–30. 10.1001/jamaophthalmol.2017.1091.

14. Joachimsen L, Farassat N, Bleul T, Böhringer D, Lagrèze WA, Reich M. Side effects of topical atropine 0.05% compared to 0.01% for myopia control in German school children: a pilot study. Int Ophthalmol. 2021;41:2001–8. 10.1007/s10792-021-01755-8.

15. Yam JC, Jiang Y, Tang SM, Law AKP, Chan JJ, Wong E, et al. Low-Concentration Atropine for Myopia Progression (LAMP) Study. Ophthalmology. 2019;126:113–24. 10.1016/j.ophtha.2018.05.029.

16. Loughman J, Kobia-Acquah E, Lingham G, Butler J, Loskutova E, Mackey DA, et al. Myopia outcome study of atropine in children: Two-year result of daily 0.01% atropine in a European population. Acta Ophthalmol (Copenh). 2024;102:e245–56. 10.1111/aos.15761.

17. Salazar M, Shimada K, Patil PN. Iris pigmentation and atropine mydriasis. J Pharmacol Exp Ther. 1976;197:79–88.

18. Myles W, Dunlop C, McFadden SA. The Effect of Long-Term Low-Dose Atropine on Refractive Progression in Myopic Australian School Children. J Clin Med. 2021;10:1444. 10.3390/jcm10071444.

19. Loughman J, Flitcroft DI. The acceptability and visual impact of 0.01% atropine in a Caucasian population. Br J Ophthalmol. 2016;100:1525–9. 10.1136/bjophthalmol-2015-307861.

20. Tideman JWL, Polling JR, Vingerling JR, Jaddoe VWV, Williams C, Guggenheim JA, et al. Axial length growth and the risk of developing myopia in European children. Acta Ophthalmol (Copenh). 2018;96:301–9. 10.1111/aos.13603.

21. Chow AHY, Mungalsingh MA, Thai D, Selimos Z, Yan SK, Xu H, et al. Suitability of multifunction devices Myah and Myopia Master for monitoring myopia progression in children and adults. Ophthalmic Physiol Opt. 2024;44:1017–30. 10.1111/opo.13332.

22. Seddon JM, Sahagian CR, Glynn RJ, Sperduto RD, Gragoudas ES. Evaluation of an iris color classification system. Invest Ophthalmol Vis Sci. 1990;31:1592–8.

23. Li FF, Zhang Y, Zhang X, Yip BHK, Tang SM, Kam KW, et al. Age Effect on Treatment Responses to 0.05%, 0.025%, and 0.01% Atropine: Low-Concentration Atropine for Myopia Progression Study. Ophthalmology. 2021;128:1180–7. 10.1016/j.ophtha.2020.12.036.

24. Moore M, Lingham G, Flitcroft DI, Loughman J. Myopia progression patterns among paediatric patients in a clinical setting. Ophthalmic Physiol Opt. 2024;44:258–69. 10.1111/opo.13259.

25. Cooper J, Eisenberg N, Schulman E, Wang FM. Maximum Atropine Dose Without Clinical Signs or Symptoms. Optom Vis Sci. 2013;90:1467. 10.1097/OPX.0000000000000037.

26. Chen Y, Li M, Shang X, Li G, Zhao Z, Li P, et al. Early changes in choroidal thickness and ocular biometry in predicting who will achieve full myopia control with repeated low-level red light therapy. Photodiagnosis Photodyn Ther. 2025;54:104672. 10.1016/j.pdpdt.2025.104672.

27. Zhang X, Liang J, Liu W, Weng Y, Chen C, Jiang H, et al. Nomogram to predict the axial elongation with orthokeratology: A 6-year follow up study. J Optom. 2025;:100590. 10.1016/j.optom.2025.100590.

